# 3D imaging of neuronal inclusions and protein aggregates in human neurodegeneration by multiscale X-ray phase-contrast tomography

**DOI:** 10.1101/2024.03.26.24304193

**Authors:** Jakob Reichmann, Jonas Franz, Marina Eckermann, Katja Schulz, Brit Mollenhauer, Christine Stadelmann, Tim Salditt

## Abstract

This study leverages X-ray phase-contrast tomography for detailed analysis of neurodegenerative diseases focusing on the 3D visualization and quantification of neuropathological features within fixed human postmortem tissue. X-ray phase-contrast tomography with synchrotron radiation offers micrometer and even sub-micron resolution, enabling us to examine intra- and extraneuronal aggregates and inclusions such as Lewy bodies, granulovacuolar degeneration, Hirano bodies, neurofibrillary tangles, ***β***-amyloid plaques and vascular amyloid deposits in three dimensions (3D). In the reconstructions, we identified the highest electron densities in Hirano bodies and Lewy bodies while neurofibrillary tangles exhibit only slight alterations in X-ray phase-contrast tomography contrast. Using cutting edge high-resolution X-ray synchrotron beamlines we are now able to even detect subcellular differences of electron densities found in granulovacuolar degeneration. Small scale inhomogeneities of the electron density are also detected in Lewy bodies potentially relating to inclusions of organelles. Additionally, we reveal a peculiar 3D geometry of Hirano bodies and demonstrate the co-occurence with granulovacuolar degeneration in the same neuron. Utilizing X-ray phase-contrast tomography in a complementary fashion to traditional technologies, a quantitative, systematic and disease-overarching understanding of inclusions and aggregates in neurodegeneration can be achieved.

## 1 Introduction

Neurodegenerative diseases are characterized by specific cellular and extra-cellular protein aggregates. Studying their three-dimensional (3D) subcellular localization and their structural composition is crucial to gain further insights into the underlying pathological processes. Alzheimer’s disease (AD) and Parkinson’s disease (PD) are the most prevalent neurodegenerative diseases, and both are characterized by intraneuronal aggregates consisting of hyperphosphorylated tau in AD and *α*-Synuclein in PD. To date, the comprehensive visualization and quantitative assessment of these neuronal aggregates, alongside extracellular *β*-amyloid plaques (*β*-APs) or co-occurring cerebral amyloid angiopathy (CAA) within a three-dimensional histological context has remained technologically challenging. For AD as well as PD but also for rarer diseases, e.g., progressive supranuclear palsy, a description of stages and of spreading of protein aggregates across the brain is in use to diagnose and estimate the neuropathological involvement [1–9]. These staging systems have to take into account the complex and folded 3D architecture of the brain with its fiber connections but usually rely on sliced two dimensional planes. In addition, the frequent coexistence of neurodegenerative pathologies in approximately 50% of either AD or PD cases underscores the need for a method enabling a quantitative 3D assessment of pathological aggregates and inclusions at subcellular resolution with comparable quality to conventional histology [1, 10]. This gap between demand and capability of 3D histological imaging has recently been narrowed by X-ray phase-contrast tomography (XPCT). As a non-destructive X-ray technique it offers high penetration, scalable resolution, and sufficient contrast for unstained native, liquid- or paraffin-embedded tissue [11–13]. Based on high spatial coherence of synchrotron radiation (SR) and even laboratory *µ*-focus sources, XPCT exploits phase contrast arising from free space wave propagation, and applicability for studies of neurodegenerative diseases has been demonstrated both for animal models as well as for human tissue, from autopsy or biopsy. For AD models in mice, for example, XPCT allowed the quantification of cellular aging [14] and the assessment of plaque morphology [15, 16] in mouse cerebellum [17, 18]. Further, XPCT was used to track neuronal loss, blood-brain barrier damage, and inflammatory cell infiltration in an experimental autoimmune encephalomyelitis (EAE) model [19–21]. Beyond animal models, investigation of biopsies of human nervous tissue by XPCT [22] - also denoted as virtual histology - was used in [23] to resolve sub-*µm* structures in the cerebellum, with notable changes in the cytoarchitecture observed for multiple sclerosis [24]. In [25], correlative imaging of XPCT and conventional histology was used to investigate AD-related pathologies of the hippocampal cornu ammonis 1 (CA1) region, further investigated in [26], where we found an unexpected chromatin compaction of granule cells of the dentate gyrus. On a larger scale, imaging of an entire liquid-embedded human brain was recently demonstrated, with the possibility to zoom in at certain areas of interest with voxel sizes down to 1 *µm* [27].

In this work, we implemented a multi-scale XPCT approach combining parallel and cone beam illumination with highly coherent 3rd and 4th generation synchrotron beams to cover a wide range of scales and to achieve high quality reconstruction of human CNS tissue, based on optimized optics, phase retrieval and reconstruction. We started out with scanning of larger volumes of paraffin-embedded tissue, fully compatible with conventional neuropathology workflows, to capture regions of interest identified, e.g. by immunohistochemistry. High-resolution zoom tomography was then used to study these areas in their native three-dimensional context, focusing on intraneuronal (e.g., Lewy bodies (LBs), granulovacuolar degeneration (GvD), Hirano bodies (HBs), neurofibrillary tangles (NFTs) and extraneuronal aggregates (*β*-amyloid plaques (*β*-APs) and vascular amyloid deposits). In a label-free approach exploiting the contrast given by varying electron densities in human tissue, we identified hallmark features of neurodegenerative pathologies, enabling both qualitative and quantitative analyses. This technique allows for three-dimensional exploration and comparison of electron densities across different pathologies, with the potential to enable insights into their generation and evolution. Further, having the 3D reconstructions obtained by non-destructive XPCT at hand, we then carried out proof-of-concept correlative immunopathological investigations. Thereby, we are able to compare electron densities of hallmark pathologies using the unique contrast mechanism of XPCT based on electron density variations in tissue. Note that for biological tissues, the electron density is in good approximation proportional to mass density. Hence, the 3D reconstructions provide important constraints for modeling biomolecular packing in aggregates and inclusions.

## 2 Results

Postmortem formalin fixed and paraffin embedded (FFPE) brain tissue from patients with neuropathologically confirmed AD (n=2), PD (n=1) and CAA (n=1) was selected for synchrotron radiation (SR) measurements. Regions of interest were identified by co-immunohistochemistry for hyperphosphorylated tau and *β*-amyloid in the CA1 region for AD patients. Temporal isocortex was assessed in CAA, whereas substantia nigra was examined in the PD patient (for patient details, see Table 1). Full 3D tomograms of tissue punches of 1 mm diameter from the respective regions of interest (see Fig.1 for an overview of the experimental approach) were recorded using two different configurations: the SR1-setup, a parallel beam configuration at DESY, Hamburg, for imaging overviews, and the SR2-setup, a cone beam configuration at ESRF, Grenoble, for high-resolution investigations (see methods and supplementary information for detailed experimental setup).

**Fig. 1:**
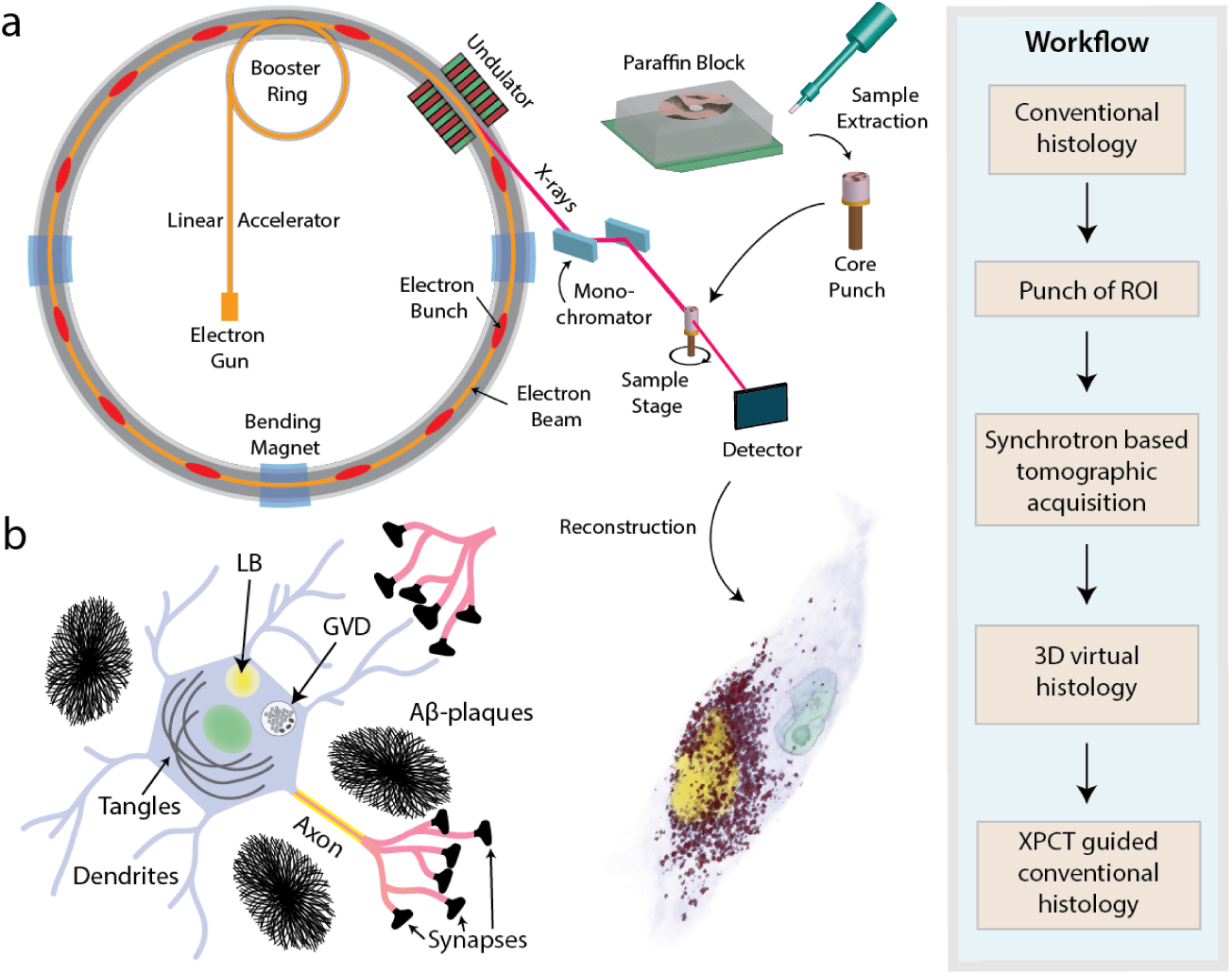
Experimental approach. **a** Sample preparation, including tissue preparation, fixation, dehydration and paraffin wax infiltration and subsequent extraction of small tissue punches. **b** Illustration of selected intra- and extra-cellular neurodegenerative pathologies and biomarkers.

**Table 1:** Patient characteristics. AD = Alzheimer’s disease, PD = Parkinson’s disease, PART= primary age related tauopathy, CAA = cerebral amyloid angiopathy, CA1 = hippocampal cornu ammonis 1, CTX = cortex, SN = substantia nigra

| Patient | Diagnosis | Age | Sex | Region Of Interest |
| --- | --- | --- | --- | --- |
| # 1 | AD (A3B3C3) | 61-65 | male | CA1 |
| # 2 | AD (A3B3C3) | 91-95 | male | CA1 |
| # 3 | PD | 76-80 | male | SN |
| # 4 | PART and CAA | 86-90 | male | CTX |

### 2.1 Intraneuronal protein aggregates and vacuoles

#### Lewy bodies in the substantia nigra closely associate with neuromelanin granules

LBs are the classical histomorphological hallmark of PD and show different morphologies depending on the neuronal subtype affected. Whereas LBs mainly consist of fibrillary a-Synuclein, other proteins as well as membranous organelles are embedded in the globular structure mostly presenting with a dense halo and a pale rim in classical histology [28, 29] . In the substantia nigra, locus coeruleus, dorsal nucleus N. V and other pigmented nuclei, LBs are in close spatial relationship with neuromelanin which is absent in most other mammals and hypothesized to facilitate LB formation [30–32]. Neuromelanin was furthermore reported to play a pivotal role in neuroinflammatory processes during progression of PD [33]. In the 3D reconstruction (Fig. 2**a**), the layered substructure of the LB is easily discernible and reveals a dense homogeneous core with a less intense surrounding shell. Also, we found that LBs in the substantia nigra are embedded in the cluster of neuromelanin granules (NMGs).

**Fig. 2:**
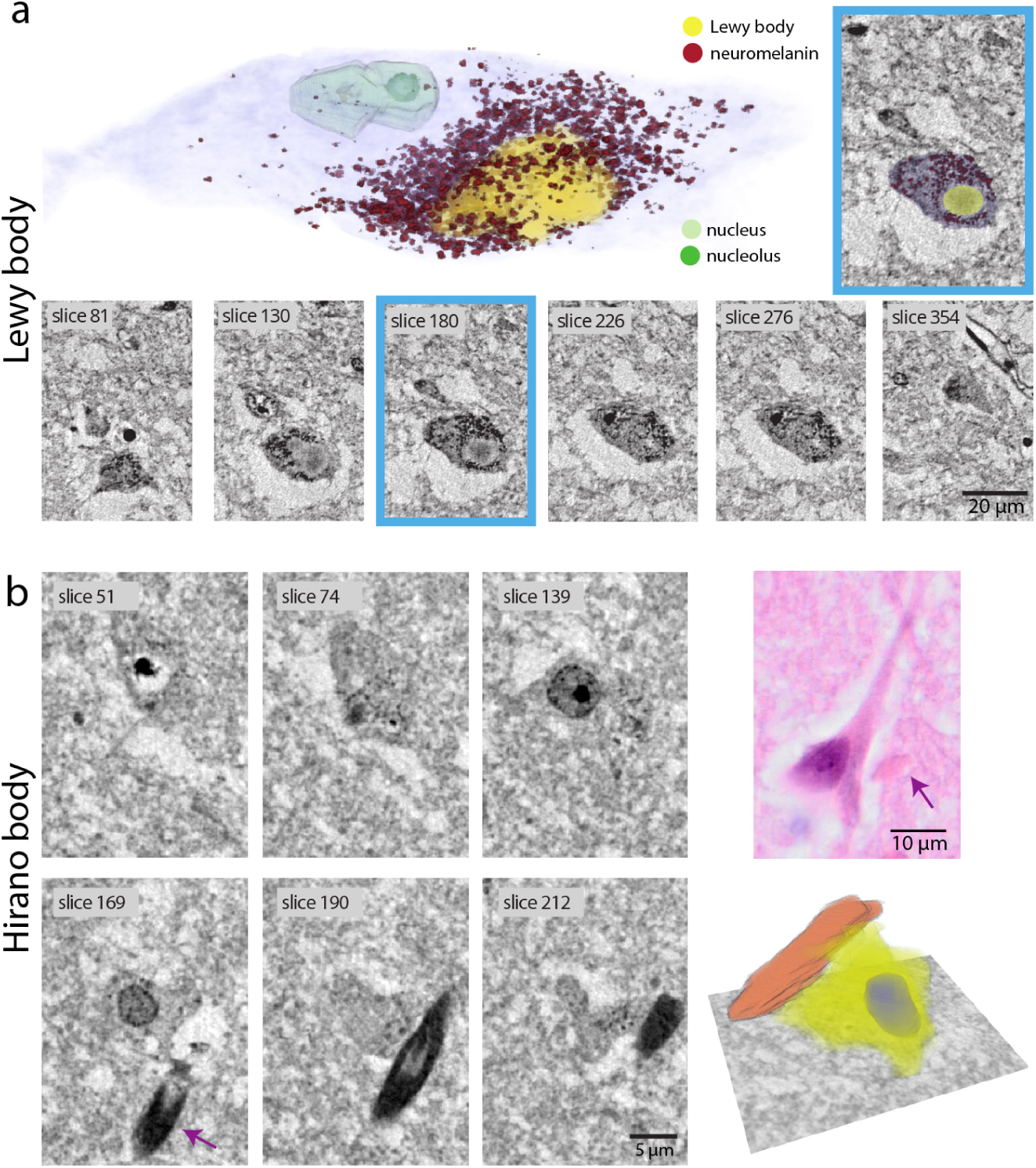
Lewy bodies (LBs) with neuromelanin granules and Hirano bodies (HBs) visualized in synchrotron *µ*CT scans. **a** Virtual section through the reconstructed volume of a dopaminergic neuron (blue: neuron surface, yellow: LB, red: neuromelanin granules, light green: nucleus, dark green: nucleolus) recorded with the cone beam SR2-setup and exemplary virtual serial slices of a LB. **b** 3D reconstruction of a HB from an AD patient CA1 region with serial sections (SR2-setup) and a HB in HE staining from the same tissue block.

#### 3D arrangement of Hirano bodies and co-occurrence with granulovacuolar degeneration

HBs are eosinophilic rod-shaped structures which mostly occur in large pyramidal neurons of the subiculum and CA1 sector in patients with AD. Their origin and role in AD pathophysiology remain largely elusive although their composition of actin filaments suggests a relation to cytoskeletal disruption. Light microscopic and EM studies suggest that HBs extend beyond the projected neuronal surface presumably leading to a cell membrane protuberance. Fig. 2**b** highlights the ability of XPCT to capture the 3D structure of HBs in human hippocampal tissue. Remarkably, HBs exhibit a pronounced contrast in XPCT compared to their typically rather inconspicuous appearance in H&E (Fig. 2**b**). The representation in Fig. 2**b** shows the 3D orientation of a HB (orange) with respect to the rest of the neuronal cell body (yellow) and the cellular nucleus (blue), demonstrating the lack of respect for the classical neuronal shape. This first human pathology based 3D representation demonstrates the relationship of the HB to the soma of the nerve cell but also shows its disruptive morphology beyond usual nerve cell borders. We also demonstrate here that HBs co-occur with granulovacuolar degeneration in the same neuron (Fig. 2**b**).

#### Granulovacuolar degeneration fills large parts of the affected neuron

GvD is a characteristic intraneuronal pathology predominantly occurring in the subiculum and CA1 region in patients with AD characterized by a grain-like basophilic core surrounded by an optically empty vacuole. The presence in brain regions particularly vulnerable to AD pathology, its correlation with NFT density and frequent occurrence in dysmorphic neurons suggest a close relationship to neurodegeneration. Also, GvD correlates with clinical dementia [7]. GvD has been proposed to reflect increased neuronal autophagy [34, 35]. Here, the 3D reconstruction enabled by XPCT allowed us to study how much of an affected neuron is occupied by GvD. Fig. 3**a** shows an in part shrunken, but on the other hand bulging neuron from the CA1 hippocampal region of an AD patient overcrowded with GvD. The high resolution, contrast and signal-to-noise ratio allow for a segmentation of individual granules and their surrounding vacuoles (Fig. 3 (**a**)). These annotations reveal a large assembly of vacuoles and granules. It appears as if vacuoles in part merge which has recently been reported in an animal model of GvD using STED-microscopy [36]. GvD seems to bulge the neuron asymmetrically. Note that GvD was also detectable in the large field of view (FOV) reconstruction obtained from the SR1-setup (supplementary information Fig. 6).

**Fig. 3:**
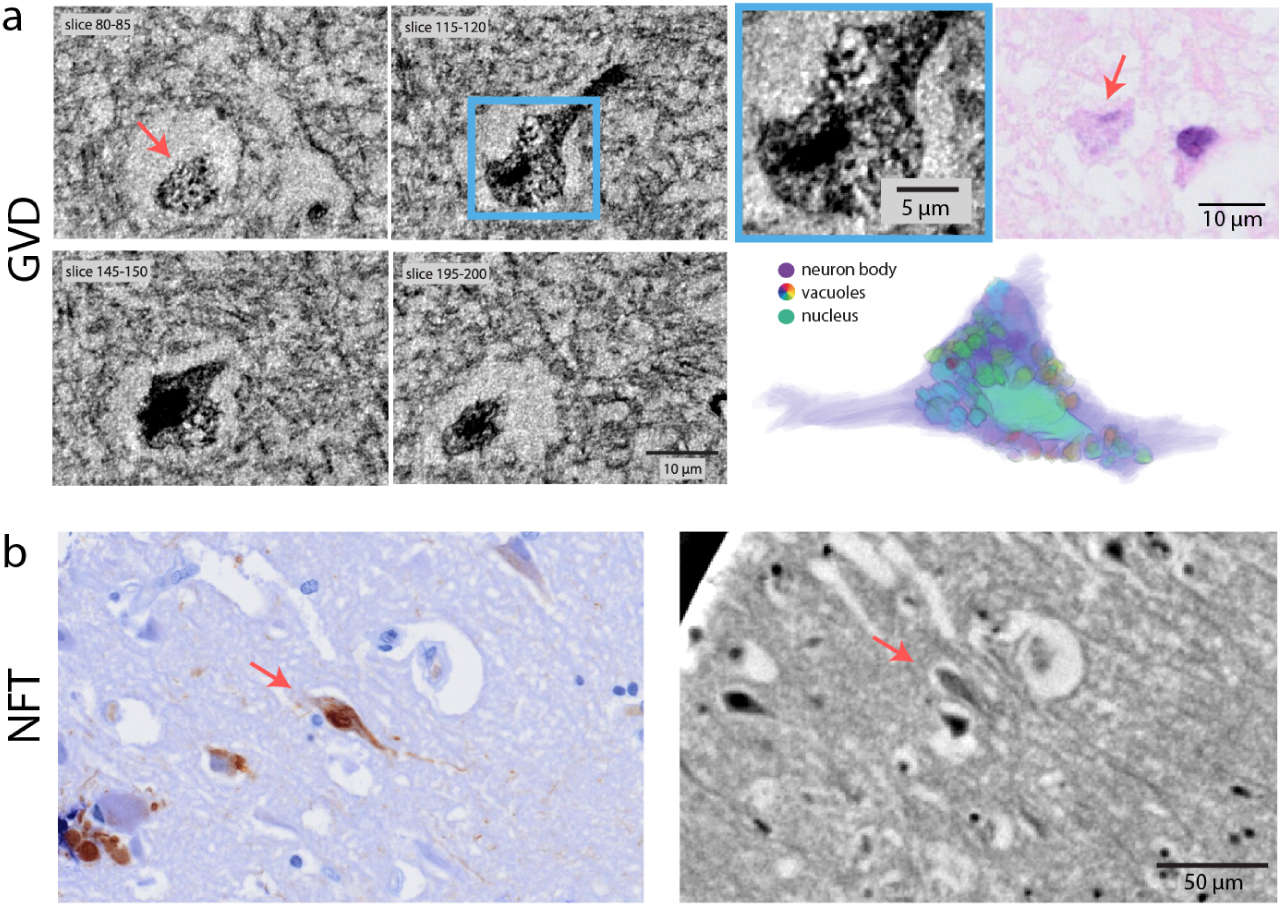
Granulovacuolar degeneration (GvD) and neurofibrillary tangles (NFTs) assessed by synchrotron *µ*CT scans. **a** 3D rendering of manually segmented granulo-vacuoles from a CA1 neuron of an AD patient’s hippocampus with virtual serial sections of GvD (maximum intensity projections, SR2-setup) and correlative histology (H&E). **b** Tangle-bearing neurons identified by correlative immunohistochemistry (AT8) and corresponding XPCT scan (SR1-setup).

#### High resolution XPCT reveals similar contrast in neurons with or without neurofibrillary tangles

NFTs consist of hyperphosphorylated tau forming a helical and barely soluble structure. Neuropathological Braak staging of AD reflects an increasing presence of NFT-bearing neurons from entorhinal to limbic to neocortical brain regions. Interestingly, the intracellular tau-aggregates were difficult to identify in XPCT, in particular if compared to the easily discernable HBs or LBs. To ensure the proper identification of individual neurons with NFTs in XPCT, we optimized the correlative immunohistochemical (IHC) analysis and applied a manual image registration approach using landmarks such as blood vessels (see supplementary information). This technique allowed us to identify tangle-bearing AT8-positive neurons visible in both the IHC-stained slice as well as in XPCT, thus enabling, e.g., comparative electron density measurements in tangle-bearing and non tangle-bearing neurons. Of note, only slight compositional heterogeneities of electron densities in the soma of affected neurons were observed (Fig. 3**b**).

### 2.2 Detection of *β*-amyloid aggregation

Aside from intraneuronal aggregates, amyloid-*β* species, which are cleaved extracellular peptides, have been central to neurodegenerative research for decades and are currently a molecular target for emerging therapies [37]. Though *β*-APs and associated microglia activation can be measured using PET, current clinical CT scanners lack the capability to visualize different plaque types or to detect amyloid deposits in arterioles, veins, or capillaries in CAA. Our approach first involved a detailed characterization of *β*-APs using the cone-beam high-resolution SR2-setup. Secondly, the feasibility to investigate CAA with the parallel beam SR1-setup was demonstrated, which is important because larger tissue volume throughput can be achieved in this setting, which is required in order to search for these pathologies.

#### Dense core of **β**-amyloid plaques yields higher **µ**CT contrast

*β*-APs are heterogeneous extracellular protein deposits characterized by varying *β*-amyloid/protein densities, different 3D structures and distinct responses of the surrounding CNS microenvironment. Their presence is an indispensable feature for AD diagnosis. In our study, even the parallel beamline of the SR1-setup allowed to delineate the comparatively dense protein core of *β*-APs. Here, an effective pixel size of 650 nm enabled the detection and analysis of cored plaques in larger tissue volumes. The improved resolution offered by the SR2-setup with a cone beam even allowed to identify the shell of a cored plaque with good contrast compared to the underlying glial matrix, as visualized in Fig. 4**a**. Employing the correlation workflow outlined above, IHC images of stained *β*-APs (using the 6E10 antibody) were juxtaposed to the corresponding XPCT measurements. Due to their size, the same amyloid plaque could easily be identified in both measurements. The second row of Fig. 4**a** displays cored plaques measured in the SR1 beamline, clearly identified by correlative IHC, and their 3D distribution. However, diffuse plaques were still difficult to detect by XPCT even with the improved resolution of the SR2-setup since they only showed a slight compaction of the texture of the tissue.

**Fig. 4:**
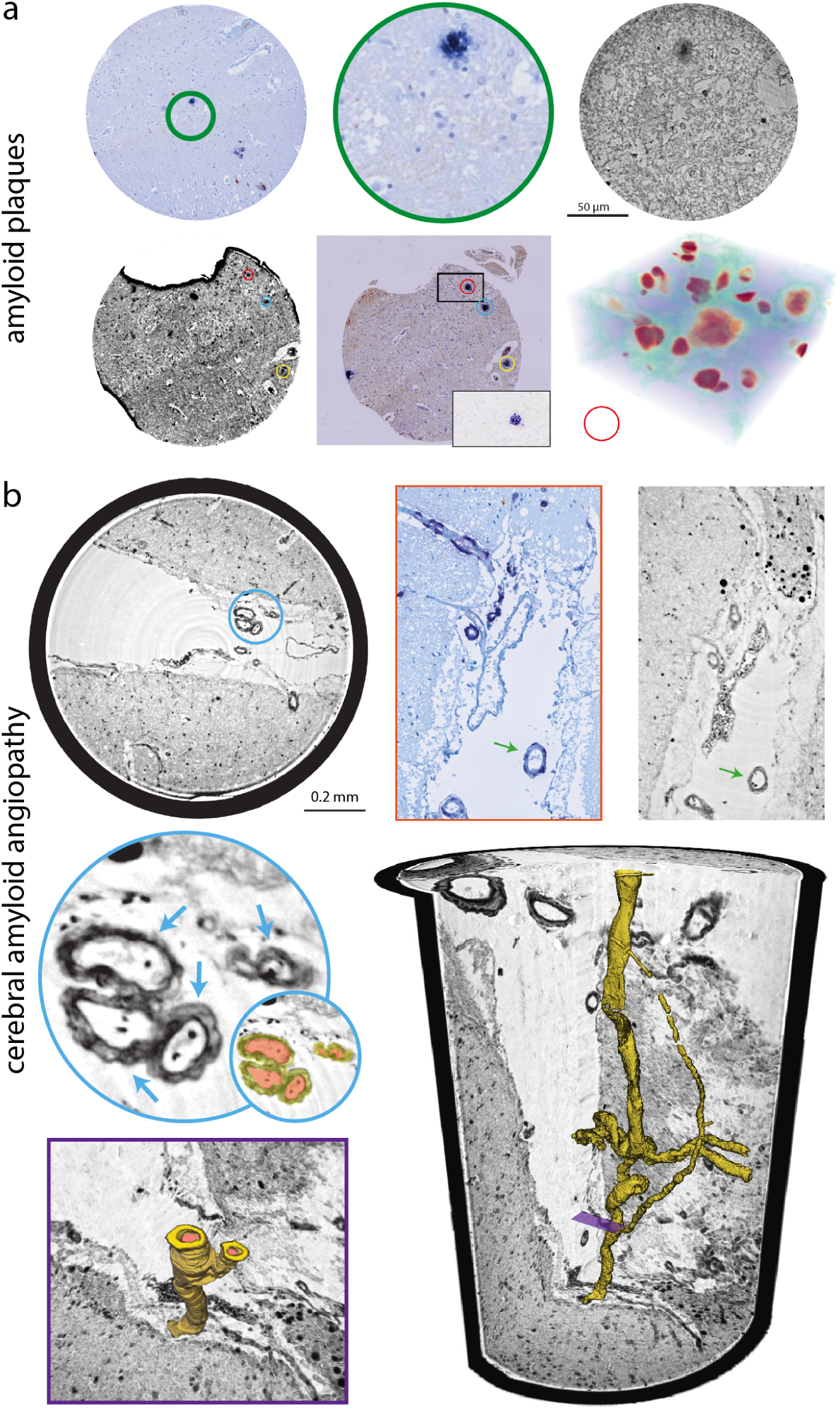
*β*-amyloid plaques (*β*-APs) and cerebral amyloid angiopathy (CAA) visualized by XPCT. **a** Correlative IHC of *β*-APs (6E10, FastBlue) and high resolution XPCT (SR2-setup). Second row: low resolution XPCT with correlative IHC (6E10) and three dimensional representation of surrounding nuclei, blood vessels and *β*-APs. **b** CAA with affected meningeal blood vessels recorded with the SR1-setup. Shown is a circular section through the 3D volume (lower right corner) with a magnified group of three blood vessels (blue) and a magnified part of the 3D rendered image (purple). Correlative IHC (red) of the affected meningeal blood vessel (green arrow) confirmed vascular amyloid deposits.

**Fig. 5:**
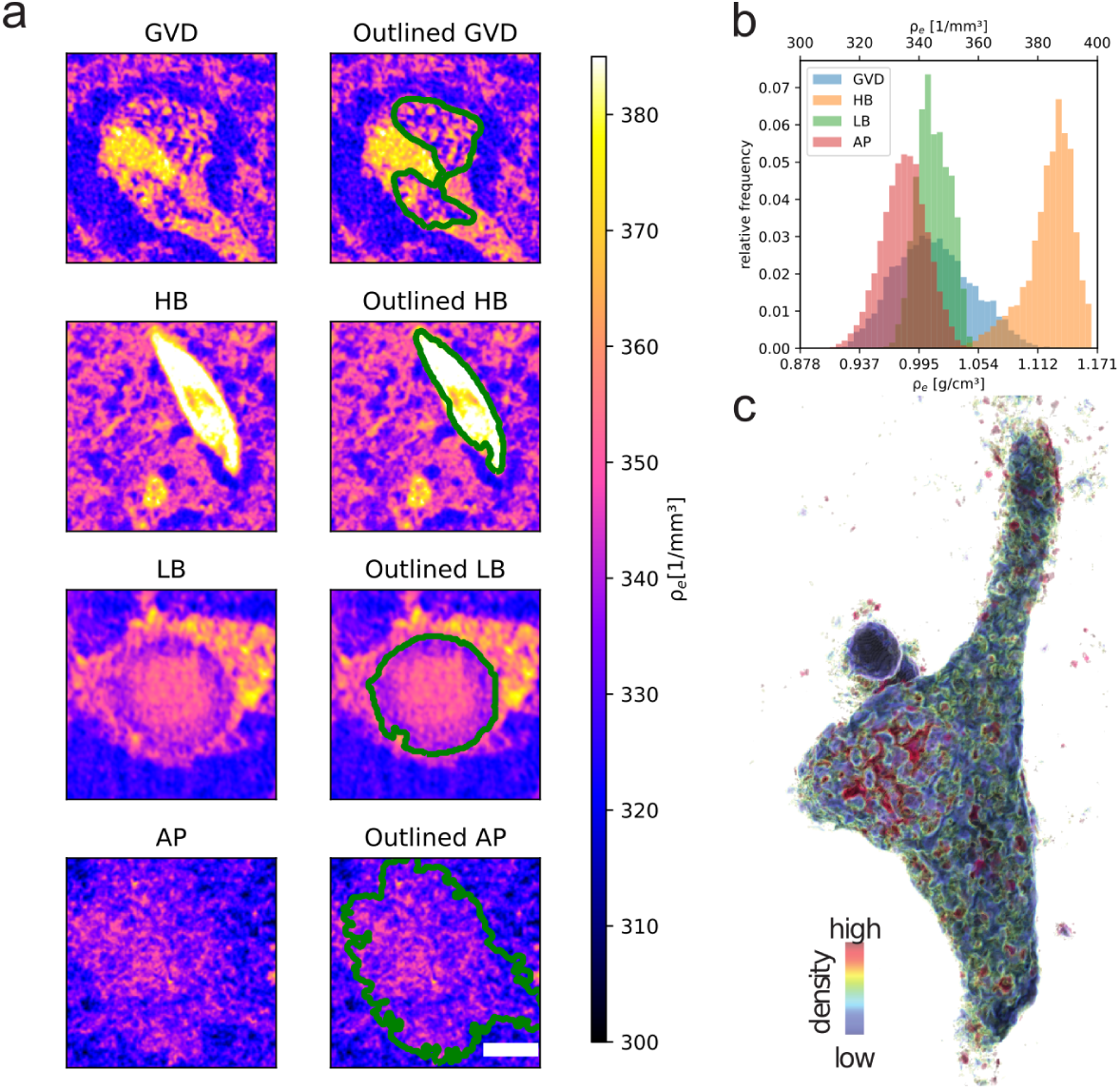
Electron densities of human neurodegenerative pathologies measured by synchrotron XPCT scans. **a** showcases cross-sectional calculated electron densities of neurodegenerative pathologies, with each row representing a different pathology (scalebar 5 *µ*m). The right images feature the same section overlaid with a green ROI delineating the electron density measurement for **b**. Panel **b** presents the electron density histograms for GvD, HBs, LBs, and *β*-APs, illustrating the distribution of electron densities within each pathological feature. Panel **c** depicts a high-resolution calculated electron density image of a neuron with GvD, color-coded to represent electron density levels from high (red) to low (blue).

#### Cerebral amyloid angiopathy

CAA may accompany AD, but may also occur independently. Characteristically, vessel wall structures are disrupted by ß-amyloid deposits leading to vascular fragility and bleeding, and more rarely, vasculitis. As for extracellular plaques, the amyloid deposits in CAA were well detectable by XPCT, also in the SR1-setup, thus enabling a multi-scale approach with larger volume throughput. Applying correlative IHC it became clear that affected vessel walls showed slightly increased contrast, see Fig. 4**b** (red). The 3D reconstruction of one exemplary affected small meningeal arteriole demonstrates the varying vessel diameter, vessel branching, and the extent of amyloid deposition (Fig. 4**b** lower right corner).

### 2.3 Quantitative comparison of electron densities obtained by XPCT

Building on the unique contrast mechanism of XPCT to delve into hallmark features of human neurodegenerative diseases, the electron density of protein aggregates and GvD was assessed. Unlike conventional histology and IHC which rely on staining-specific amplification methods, the image formation of XPCT is directly linked to the electron density distribution. Using standardized acquisition and analysis, the electron density can be calculated from the phase shifts obtained by phase retrieval. Therefore, image gray values can be regarded as quantitative measurements of the local electron density. While this relationship can be more difficult to establish when absolute numbers are required, it is easier and more robust to provide analysis on local differences in electron density. Here the paraffin embedding provides a well suited reference to ’calibrate’ electron density. Since electron density (or correspondingly the mass density) effectively mirrors biological parameters such as fibril density or the presence of metal ions, it can provide important additional information on a given pathology. Of note, the standardized image acquisition process allows a direct comparison of neuronal and extracellular protein aggregates and structures, even across different diseases (for details on electron density calculation, see supplementary information).

Electron density assessment revealed small inhomogeneities on the scale of 2-3 μm within the LB, predominantly in the so-called ”halo”, compatible with the notion that organelles may be encased in the fibrillary *α*-synuclein network (see 5**a** and Fig.7 in[28]). Notably, the calculated quantitative electron density revealed a circular arrangement of these hypodense, potentially lysosomal organelles at the outer part of the LB, providing insights into the subcellular composition of this aggregate.

Using this approach, we could assign to HBs electron densities comparable to those of neuromelanin granules (see 5**a** + **b**), underscoring the utility of XPCT in differentiating between various intracellular aggregates based on their electron density profiles. GvD, in contrast, and as expected, did not display a homogeneous distribution of electron densities. However, we also could not ascertain a clearly bimodal distribution. This is suggestive of vacuoles showing a slowly increasing density towards their central ”grain”. Lastly, consistent with its imaging behaviour, the extracellular amyloid plaque seemed to be composed rather loosely, and only the core showed a slight increase in electron density.

## 3 Discussion

Protein aggregates are a central hallmark of numerous neurodegenerative diseases instructing genetic and pathogenetic research; however, their formation, cytoplasmic embedding, and role for neuronal toxicity are not resolved. In this work, we leveraged XPCT of paraffin-embedded autopsy tissue combined with correlative immunohistology to identify and compare disease-defining pathological protein aggregates and inclusions in their full 3D and at sub-cellular scale. Our studies revealed a close spatial relationship of neurodegenerative protein aggregates to other intracellular structures, e.g. neuromelanin, and distinct properties with regard to electron density. While we observed a particularly high electron density in HBs, NFTs were barely visible and required correlative imaging techniques. In GvD, instead of the expected bimodal density distribution, we found a gradient of increasing electron densities towards the central ”grain”, suggesting increasing protein content. In summary, our work combines advanced X-ray imaging techniques and correlative immunohistology in neurodegenerative diseases demonstrating its capability for 3D assessment of protein aggregates and inclusions on the tissue and subcellular levels.

While the general compatibility of XPCT and FFPE samples has been previously established [25, 26, 38], we demonstrate here that improvements in resolution and contrast, facilitated by the specificities of the SR2-setup (see below), are sufficient to identify and to quantify also subcellular neuronal pathologies, such as LB, GvD, and HB. To our knowledge, some of these structures have never or only rarely been imaged in 3D before.

The size and orientation of protein aggregates relative to the cellular body and nucleus may have significant implications, offering valuable insights into their development within the cell. For instance, the peculiar orientation of HB revealed through 3D virtual histology by XPCT (see Fig. 2**b**) may give indications with regard to its mechanism of formation. Likewise, the 3D association of neuromelanin around LB observed here (see Fig. 2**a**) is noteworthy and increasing the evidence for an association of fibrillary alpha-synuclein with neuromelanin pigments derived from extensive two dimensional histological studies [30]. The usual size of LB ranges from 5 to 25 *µ*m in diameter with a dense eosinophilic core of filamentous and granular material surrounded by radially oriented filaments [39, 40]. This core and the filamentous shell are likely to be reflected in the XPCT reconstruction in form of elevated levels of electron density by XPCT. There is also recent evidence using confocal as well as super-resolution stimulated emission depletion (STED)-microscopy combined with electron microscopy and tomography that dense lysosomal structures and a shell of distorted mitochondria surround some of the LB inclusions (see Fig. 7 of [28]). In the present study we were able to even identify these LB substructures, owing to the superior resolution provided by the SR2-setup.

While the 3D inspection alone allowed us to better understand the nature of the aggregates, their distribution in the tissue and their relation to other intracellular structures, the quantitative analysis of XPCT contrast values revealed significant differences in electron densities between subcellular aggregates.

While the 3D inspection alone allowed us to better understand the nature of the aggregates, their distribution in the tissue and their relation to other intra-cellular structures, the quantitative analysis of XPCT contrast values revealed significant differences in electron densities between subcellular aggregates. By far the highest electron density was found in HBs. This finding is in line with previous electron microscopy studies using quick-freeze deep-etch technology where HBs were demonstrated to consist of intracellular and densely packed fibrillary aggregates [41]. Compared to HBs the distribution of electron densities in LBs is lower, hinting towards less densely aggregated proteins or an inclusion of elements with higher electron density like metals in HB. The electron density in LBs is in the range of that of the GvD granules. However, GvD exhibits a broader distribution with a lower mean value accounting also for the vacuoles. The lowest values in GvD approximately reach a density of 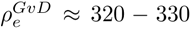 (compare to paraffin with 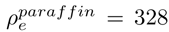). This would relate to the almost protein free vacuolar space in between granules. Note, however, that this absolute result relies on the assumed density of the paraffin wax in our analysis and the corresponding reference normalization.

Here, and to our knowledge for the first time, correlative immunohistochemistry was used to facilitate, at a single cell resolution, the identification of neurons with NFTs in XPCT. Even though the identification of tangle-bearing neurons in the investigated samples was not yet possible by XPCT alone, the highly precise measurements and definitive identification of tangles through well established methods allowed us to draw pertinent conclusions. Of note, and compared to HBs and LBs, tangles do not lead to a homogeneous increase in the electron density of affected neuronal cytoplasms. Relating this finding to existing electron microscopy and STED-microscopy studies thus suggests that individual NFTs are surrounded by numerous subcellular organelles and do not densely fill up the neuron [42–44]. XPCT may therefore serve as a complementary method to highly advanced cryo-EM resolving the atomic structure of protein aggregates, for example, of tau filaments in AD [45]. We can hypothesize from our studies that the pathologic effect of tangles may be mediated more by a derangement of the cytoskeleton, especially the microtubules, obviously leading to various cellular dysfunctions, rather than by completely replacing existing subcellular organelles as observed in HBs. Our study also raises the question if extended XPCT methods implementing X-ray contrast agents or increased resolution will be able to identify pathological tau filaments. Note that the resolution of XPCT has not yet reached its fundamental limits, and can be expected to be further increased by ongoing instrumental improvements.

Also for *β*-APs, correlative IHC further improved our detection sensitivity as demonstrated by high resolution images of the SR2-setup (see Fig. 4**a**). *β*-APs were recently identified by XPCT in unstained human autopsy brain tissue by Chourrout et al. [15], reporting varying contrasts in mice and humans supposedly depending on the degree of calcium accumulation, in line with findings by Toepperwien et al. where mineralized plaques could be clearly observed even with in-house *µ*CT instrumentation [23]. Our electron density measurements revealed that the core of *β*-APs showed a significantly higher electron density while the less dense shell is only barely visible even in the high resolution SR2-setup. One may wonder why *β*-APs or NFTs are difficult to detect in XPCT. Since for FFPE tissue, contrast is generated by the difference with respect to the embedding paraffin matrix, this can happen for organelles or structures which happen to exhibit similar density as the matrix. As a solution, contrast variation by different embedding media could be employed [11].

Not only but also in the light of amyloid therapy-related adverse effects a better understanding of vascular amyloid deposition and microhemorrhage is important (Salloway 2021, Cummings 2023). CAA and *β*-amyloid related angiitis (ABRA) represent a significant risk factor for intracerebral micro- and macrohemorhages. Thus, a 3D visualization of microvascular changes may help to scrutinize effects of amyloid depositions in blood vessels also as an effect of amyloid targeting therapies. The highly precise *µ*-CT strategy developed in this study (Fig. 4**b**) demonstrates a new scale for the identification of even minor blood vessel changes invisible to angiography and for investigations of small vessel branching.

While the voxel size and instrumental resolution (as determined for high contrast objects) are clearly sufficient to image the targeted pathologies at subcellular scale, the reconstruction of the FFPE tissue still exhibits substantial noise. In fact, as outlaid above, it is the low contrast of specific features such as unmineralized *β*-APs which limits the 3D structural information gained rather than the instrumental resolution. For the SR2-setup in particular, 50 nm (half period) resolution has been achieved in inorganic samples with high contrast or in metalized biological specimens [13, 46]. In order to reduce noise and to increase contrast in the present unlabeled FFPE tissues, one can either increase dose or decrease the photon energy *E*. Since the phase shift Δ*ϕ* per resolution element scales with *E^−^*^1^ (away from absorption edges), the contrast increases accordingly. Note that the diameter of the biopsy punches would clearly allow a reduction of *E* by at least a factor of two.

Future extension of this work may also include recently developed X-ray stains [47–50] and labels to improve contrast not only for distinct structural hallmarks of neurodegeneration covered in this work, but also for the individual cellular components, e.g., axons, dendrites or synapses, or the myelin sheath which would open up a new perspective for X-ray imaging of myelin-associated diseases, such as multiple sclerosis.

In summary, our work combines X-ray phase contrast tomography and correlative immunohistology on paraffin-embedded tissue specimens of neurodegenerative diseases. This approach allows us to identify protein aggregates and other inclusion bodies and reveals their 3D distribution in tissue, relevant, e.g.. for studies of spreading of pathology. In addition, this multi-scale approach enables subcellular 3D assessment of inclusion bodies, revealing insights into their relation to subcellular organelles. Upcoming technological developments will further increase the usefulness of this technology for the study of human CNS diseases.

## 4 Methods

### Sample preparation

Human brain tissue from individuals who underwent diagnostic autopsy in the context of routine clinical care was obtained from the archives of the Department of Neuropathology UMG in accordance with UMG ethics regulations. Small brain tissue blocks were dissected from 10% formalin-fixed brain slices, dehydrated, and paraffin embedded (see [26]). One FFPE tissue block measured about 2×3×0.3 cm^3^. In total, four samples from four individuals were selected for the study, including two samples from the hippocampal CA1 region (both with immunohistochemically confirmed NFTs (see immunohistochemistry)), one sample from the substantia nigra from a PD patient and one from the temporal cortex including the leptomeninges of a patient with CAA. Regions or interest were defined on an adjacent histological section. To prepare the samples for XPCT image acquisition, cylindrical 1 mm biopsies were extracted from the paraffin blocks, inserted in a polyimide tube and placed on Huber pins. After imaging, these paraffin blocks were again embedded in paraffin and processed for further histological or immunohistochemical analysis.

### Immunohistochemistry

Immunohistochemistry was performed on 2-3 μm thick paraffin sections. Pre-treatment included hydrogen peroxide as well as formic acid (98%) (for A*β* immunhistochemistry only), blocking in 10% normal goat serum as well as heat antigen retrieval (citrate buffer, pH 6). Incubation of primary antibodies over night (tau: mouse, clone at8, Thermo Fisher Scientific, 1:100; *β*-amyloid: mouse, clone 6E10, Zytomed Systems GmbH, 1:500) was followed by secondary antibody incubation either coupled to alkaline phosphatase (polyclonal goat anti-mouse, Dako, 1:50) or to biotin (monoclonal sheep anti-mouse, GE Health-care Life Sciences, 1:100). Slides were developed using avidin-peroxidase with DAB and/or fast blue.

### Propagation-based phase-contrast imaging at the synchrotron

Synchrotron radiation allows for imaging with high coherence and brilliance and can cover multiple length scales down to sub-100 nm resolution . In this work we take advantage of a multiscale XPCT approach, using the parallel beam (SR1) configuration of the GINIX endstation installed at the P10 beamline of the PETRA III storage ring (DESY, Hamburg) [51] and the nano-imaging beamline (SR2) ID16A (ESRF, Grenoble, France). While the former is optimized for larger FOV with a parallel beam geometry, the latter is dedicated to propagation-based holographic tomography of biological samples, and operates in the hard X-ray regime (17-33.6 keV). To achieve high resolutions, the beam is focused by a pair of Kirkpatrick-Baez (KB) mirrors. Both provide quantitative phase contrast which allows to retrieve information on the electron density in the sample, making them especially useful for biomedical applications. The two configurations can be used complementarily to study biological tissue at multiple scales. Details on the setup and acquisition parameters can be found in the Supplementary Document.

- SR1: The imaging procedure involved an overview scan using a field of view (FOV) of approximately 1.5 mm. Therefore, a parallel-beam configuration was employed, enabling continuous rotation and resulting in an overall scan time of approximately 2 minutes. The acquisition of single-distance tomograms involved 3000 projections captured over a 360° rotation. To achieve a well-defined photon energy for studying the tissue, a Si(111) channel-cut monochromator was utilized, eliminating the broad band-pass limitations inherent in in-house CT systems. On the registration side, a high-resolution detection system (Optique Peter, France) with a 50 mm-thick LuAG:Ce scintillator and a 10× magnifying microscope objective [51] was coupled with the sCMOS camera pco.edge 5.5 (PCO, Germany). The camera performs with a maximum frame rate of 100 Hz, utilizing a rolling shutter and fast scan mode. This configuration yielded an effective pixel size of 0.65 *µ*m, with an approximate total exposure time of 96 s. To achieve the desired imaging conditions, certain components such as KB-mirrors, waveguide, and the fast shutter were removed from the beam path. Additionally, the beam size was adjusted to approximately 2×2 mm using an upstream slit system (refer to Fig. 1**d**).
- SR2: The nano-imaging beamline ID16A provides a highly-brilliant, low-divergent beam optimal for three dimensional high-resolution imaging of biological samples or other nanomaterials e.g. in batteries. With its multi-layer monochromator and focusing KB-mirrors, it allows for photon energies of either 17.1 or 33.6 keV and a photon flux of up to 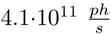. The cone beam geometry enables a magnification of the projected pixels resulting in possible effective pixel sizes *px_eff_* of less than 10 nm. The monochromaticity allows for a subsequent quantitative analysis and retrieval of sample characteristics such as electron density. Due to the strong magnification and high coherence, phase propagation can be very accurately recorded and reconstructed from the strongly holographic projections (F*≪*1). For detection, a XIMEA sCMOS based indirect imaging detector with 6144 x 6144 pixel (10 *µ*m physical pixel size) and a 10× magnifying microscope objective was used. In this experiment the projections were binned (3×3), with effective pixel sizes ranging from 90 to 140 nm. 2000 projections were recorded per scan with additional random sample displacement for every angle to correct for wavefront inhomogeneities and therefore avoiding ring artifacts [52]. Tomographic scans were acquired at four distances to account for zero crossings in the CTF phase reconstruction [53]. From the four distances, the one with the highest resolution and largest FOV is combined, resulting in an “extended FOV” of 3216^2^ pixel in the resulting tomographic slices (refer to Fig. 1**e**).

### Data processing

Projections were acquired and saved using the .tiff or .raw format. After flat field and dark image correction, operating in the holographic regime, phase retrieval was performed using either the contrast transfer function (CTF, [53, 54]), the nonlinear Tikhonov (NLT, [55]) or a Paganin-based iterative scheme [56] (see supplementary information). On the phase retrieved projections, ring removal techniques were conducted (only random displacement at SR2-setup) as well as an automatic rotation axis correction. Tomographic reconstruction was then performed by either filtered back projection (parallel beam, SR1) or the Feldkamp-Davis-Kress (FDK, [57]) algorithm. Both techniques are implemented in the ASTRA-Toolbox [58] for MATLAB and incorporated into the HolotomoToolbox [59]. Further details on the different reconstruction schemes can be found in the supplementary information.

### Segmentation and visualization of cell components

After tomographic reconstruction of the recorded, phase-retrieved, projections and visual inspection, different structures were proposed for further analysis. After selection, segmentation was conducted using seeded watershed algorithms, deep learning based techniques (webKnossos [60], scalable minds GmbH, Potsdam, Germany) or simple thresholding. Subsequently, rendering software such as NVIDIA IndeX (NVIDIA, Santa Clara, US), Avizo (Thermo Fisher Scientific, Waltham, US) and ZEISS arivis (Carl Zeiss AG, Oberkochen, Germany) was used for a three-dimensional visualization of the dataset. For additional post processing such as orthogonal views and maximum intensity projections the Fiji software was used [61]. Segmentation masks for electron density calculations were manually generated using QuPath [62]. Details on segmentation and electron density calculations can be found in the supplementary information.

## 5 Declarations

### Ethics approval and consent to participate

Brain tissue blocks of patients with neurodegenerative diseases were obtained during diagnostic autopsy and retrieved from the archives of the Institute of Neuropathology, University Medical Center Göttingen, Germany. The tissue was used in an anonymized fashion. The study was approved by the ethics committee of the University Medical Center Göttingen (22/1/19).

### Consent for publication

For the purpose of open access, the authors have applied a CC BY public copyright license to all Author Accepted Manuscripts arising from this submission.

### Data availability

Raw data were generated at ESRF and DESY. Raw data will be released and made public two years after the beamtime. All treated datasets are available from the corresponding authors on reasonable request. Exemplary datasets that support the findings of this study will be openly available in GRO.data upon publication.

## Data Availability

Raw data were generated at ESRF and DESY. Raw data will be released and made public two years after the beamtime. All treated datasets are available from the corresponding author on request. Exemplary datasets that support the findings of this study will be openly available in GRO.data upon publication.

## Competing interests

The authors declare no competing interests.

## Funding

The work was funded by the Deutsche Forschungsgemeinschaft (DFG, German Research Foundation) – Project-ID 432680300 – SFB 1456/A03 *Mathematics of Experiment* and under Germany’s Excellence Strategy - EXC 2067/1-390729940. This research was funded in part by Aligning Science Across Parkinson’s ASAP-020625 through the Michael J. Fox Foundation for Parkinson’s Research (MJFF). JF was supported by the UMG Clinician Scientist Program. JR was supported by the Hertha Sponer College (MBExC).

## Contribution statement

JR and JF contributed equally to this work. TS, CS, JF and JR conceived the experiments and analysis. JF selected and prepared the samples. JR, TS conducted the synchrotron experiments, together with ME, acting as local contact at the ID16A beamline. KS and JF performed correlative immunohistochemistry. JR performed data processing, image reconstruction and visualization. JF annotated data and performed data processing. JR, JF, and CS interpreted the results in terms of neuropathology. BM supported tissue procurement. TS, CS, JF and JR wrote the manuscript. All authors reviewed the manuscript.

## Acknowledgements

We thank Markus Osterhoff, Michael Sprung and Fabian Westermeier for their continuous support at the instrument GINIX/P10 (PETRA III, DESY). We also acknowledge assistance in visualization with NVIDIA IndeX (NVIDIA Corporation, USA). We thank Heidelinde Brodmerkel, Reńe Müller and Melina Wüstefeld for excellent technical assistance.

## Notes

### Competing Interest Statement

The authors have declared no competing interest.

### Author Declarations

Ethics approval and consent to participate} Brain tissue blocks of patients with neurodegenerative diseases were obtained during diagnostic autopsy and retrieved from the archives of the Institute of Neuropathology, University Medical Center Goettingen, Germany. The tissue was used in an anonymized fashion. The study was approved by the ethics committee of the University Medical Center Goettingen (22/1/19).

### Summary of Updates

The title was updated and a general revision of the text, in particular regarding the results and the discussion sections, was conducted. Author affiliations were updated, as well as the supplemental files.

