## Supplemental Document 1 for "3D imaging of neuronal inclusions and protein aggregates in human neurodegeneration by multiscale X-ray phase-contrast tomography"

### Experimental Setup - X-ray phase-contrast tomography

In the following, all relevant experimental settings are tabulated. Hereinafter, the parallel beam setup at the GINIX endstation (Hamburg) is referred to as synchrotron radiation 1 (SR1), while the nano-imaging beamline at the ID16A at the ESRF (Grenoble) is referred to as SR2. Both setups are illustrated in Figure 1. Note that for SR2 the $x_{01}$ distance refers to the first distance of a potential multi-distance tomographic scan. The dose deposited on the sample recording a single projection was estimated as:

$$D[Gy] = \frac{I_0 \tau E}{d \rho_m \text{hv}}, \tag{1}$$

with the photon flux $I_0$, exposure time $\tau$, photon energy $E$, attenuation length $d$, mass density $\rho_m$ and the vertical and horizontal extent of the illuminated field of view in the sample plane (FOV), $h$ and $v$, reciprocally. The photon flux on the sample was estimated as $\mathrm{I_0} \approx 5 \cdot 10^{11}$ ph/s and $\mathrm{I_0} \approx 4.1 \cdot 10^{11}$ ph/s for the parallel beam setup (SR1) and the high-resolution setup (SR2), respectively. The FOV was $1.6 \times 1.4\text{mm}^2$ (h $\times$ v) for SR1, and $(0.21 - 0.45) \times (0.21 - 0.45)$ mm$^2$ (h $\times$ v) for SR2 (first distance). For the attenuation length $d$, $3 \cdot 10^{-3}$ m (SR1) and $0.59 \cdot 10^{-3}$ m (SR2) were used as computed by Henke et al. (1993) for a representative protein and tissue composite with stochiometry $C_{50}C_{30}N_9O_{10}S_1$. The mass density of paraffin embedded samples was estimated by 0.8 g/cm$^3$, as suggested in Kaye & Laby (1995).

### Data Post-Processing

#### *Phase Retrieval and Tomographic reconstruction*

As a first step, all projections are flat- and dark field corrected, according to the scheme $\frac{\text{proj}-\text{dark}}{\text{flat}-\text{dark}}$. In propagation-based x-ray phase contrast imaging, a single projection on the detector is formed by free space propagation of the exit wave $\psi_e$ behind the object, which in projection approximation is written as

$$\psi_e(y,z) = \psi_0(y,z)\ \tau(y,z)\ , \tag{2}$$

with $\psi_0$ the illumination function and $\tau(y,z)$ the complex-valued object transmission function, accounting for the attenuation and the phase shift introduced by the object.

Table 1: Experimental parameters for the parallel beam synchrotron setup SR1. FOV denotes the field of view (horizontal and vertical), $x_{01}$ the source-to-sample distance, $x_{12}$ the sample-to-detector distance, $px_{eff}$ the effective pixel size, $F$ the Fresnel number and $D$ the dose.

| | SR1 |
|---|---|
| # projections | 3000 |
| FOV (h × v) ($mm^2$) | 1.6 × 1.4 |
| $x_{01}$ | 88 m |
| $x_{12}$ | 33 mm |
| $px_{eff}$ | 650 nm |
| Exp. time (s) | 0.045 |
| Tot. exp. time (s) | 96 |
| Energy (keV) | 8.08 |
| F | 0.083 |
| D ($\times 10^4$ Gy) | 1.63 |

The intensity recorded in the detection plane then is given by as $I = |D_x(\psi_r)|^2|$, where $D_x$ is the Fresnel propagator. As an approximative forward model in the holographic regime for weak objects, the contrast transfer function (CTF) in Fourier space is given by

$$\frac{\hat{I}_\omega(\mathbf{k}_\perp, x)}{I_0} = 2\pi\delta_D(\mathbf{k}_\perp) + 2\sin\frac{\lambda x(\mathbf{k}_\perp)^2}{4\pi}\hat{\phi}_\omega(\mathbf{k}_\perp) - 2\cos\frac{\lambda x(\mathbf{k}_\perp)^2}{4\pi}\hat{\xi}_\omega(\mathbf{k}_\perp) \ , \qquad (3)$$

where $\mathbf{k}_\perp$ represents the components of the spatial frequency vector in the plane normal to the optical axis, $\lambda$ the wavelength, while $\phi_\omega$ and $\xi_\omega$ are the Fourier transformed attenuation and phase maps. Thus, we can observe an oscillatory contrast in phase and amplitude, both along the propagation axis $x$ and in the $\mathbf{k}_\perp$plane. This implies zero crossing in the contrast transfer, resulting in missing frequencies and therefore loss of information. By recording tomograms at several different propagation distances, this can be compensated, thus filling up the reciprocal space and accounting for the missing frequencies.

To extract the encoded object information, phase retrieval algorithms are required, solving the inverse problem. For the SR1 experiments presented in this manuscript, CTF-based phase retrieval Cloetens et al. (1999) was carried out, assuming a slowly varying phase and a low-absorbing object (see CTF above), resulting in a regularized Fourier filter operation, which can be written for $N$ different propagation distances as

$$\phi(\mathbf{r}_\perp) = \mathcal{F}_\perp^{-1}\frac{\sum_{m=1}^{N}[sin\chi_m + (\beta/\delta)cos\chi_m]\mathcal{F}_\perp[I_\omega^{exp}(\mathbf{r}_\perp, x) - 1]}{\sum_{m=1}^{N} 2[sin\chi_m + (\beta/\delta)cos\chi_m]^2 + \alpha(\mathbf{k}_\perp)}, \chi = \frac{\lambda z \mathbf{k}_\perp^2}{4\pi} \ , \qquad (4)$$

Table 2: Experimental and reconstruction parameters for the high-resolution synchrotron setup SR2. # defoc. dist. denotes the number of recorded distances, FOV the field of view (horizontal and vertical), $x_{01}$ the source-to-sample distance, $x_{02}$ the source-to-detector distance, $px_{eff}$ the effective pixel size, $F$ the Fresnel number and D the dose. As for the reconstruction parameters, $lim1$ and $lim2$ denote the upper and lower cut-off frequency for the contrast transfer function and $\delta/\beta$ the ratio between refraction and attenuation in the sample.

| | Lewy-Body | Hirano Body | GVD | TE |
|---|---|---|---|---|
| # projections | 2000 | 4000 | 4000 | 2000 |
| # defoc. dist. | 4 | 4 | 3 | 4 |
| FOV (h × v) ($mm^2$) | 0.45 × 0.45 | 0.29 × 0.29 | 0.21 × 0.21 | 0.45 × 0.45 |
| $x_{01}$ (mm) | 53.1 | 34.7 | 36.5 | 53.1 |
| $x_{02}$ (m) | 1.2 | 1.2 | 1.2 | 1.2 |
| $px_{eff}$ (nm) | 140 | 90 | 90 | 140 |
| Exp. time (s) | 0.2 | 0.2 | 0.2 | 0.2 |
| Energy (keV) | 17.1 | 17.1 | 17.1 | 17.1 |
| F ($\times 10^{-4}$) | 5.4 | 2.2 | 2.2 | 5.4 |
| lim1 ($\times 10^{-3}$) | 0.1 | 0.1 | 0.1 | 0.1 |
| lim2 | 0.01 | 0.01 | 0.01 | 0.01 |
| $\delta/\beta$ | 80 | 80 | 80 | 80 |
| D ($\times 10^6$ Gy) | 4.2 | 20.1 | 38.4 | 4.2 |

with the frequency dependent regularization parameter $\alpha(\mathbf{k}_\perp)$ and $I_\omega^{exp}(\mathbf{r}_\perp, x)$ representing the experimentally measured intensity. The phase retrieval was carried out using the numerical implementation given in Lohse et al. (2020). For the SR2 data, an iterative phase retrieval scheme that extends Paganin's single distance method was used (Yu et al. (2018)). Following the phase retrieval of the projections and a ring-filter step as outlined in Münch et al. (2009), the tomographic reconstruction was carried out using either the filtered back-projection (FBP) method or the cone-beam algorithm (FDK), both of which were implemented in the ASTRA-toolbox (van Aarle et al. (2015)).

*Segmentation and visualization of features of interest*

After completing the tomographic reconstruction of the phase retrieved projections and subsequent visual inspection, a range of structures were put forth for further analysis. Depending on the particular dataset, different segmentation methods were explored. For structures with well-defined edges, seeded watershed algorithms implemented in the

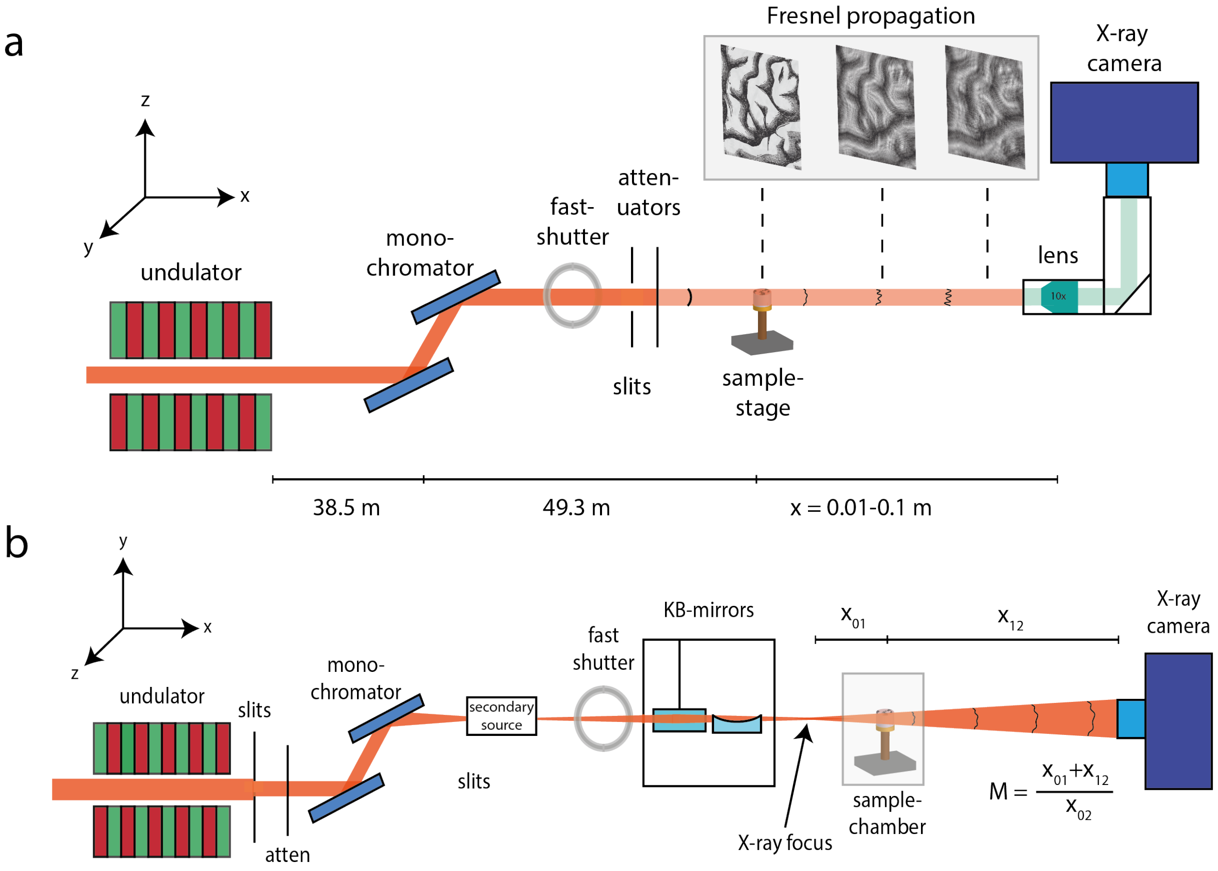


Figure 1: **a** Parallel beam setup at the GINIX instrument, P10 beamline, DESY, Hamburg (photo in supplementary document, detailed description in the text). **b** High-resolution holo-tomography setup at the ID16A beamline, ESRF, Grenoble (photo in supplementary document, detailed description in the text).

.

*Carving* option in the interactive learning and segmentation platform Ilastik (Berg et al. (2019)) was used (see Fig. 4 in main manuscript). If the contrast was sufficient, pixel classification was based on machine learning algorithms (also implemented in Ilastik), as an alternative to manual segmentation or simple thresholding which often encounters difficulties. For shape-based segmentation, machine learning and deep learning based tools implemented in the ZEISS arivis software (Carl Zeiss AG, Oberkochen, Germany) and webKnossos Boergens et al. (2017) (scalable minds GmbH, Potsdam, Germany) were applied. To this end, regions of interest (ROI) were manually annotated in a few slices, whereupon the features in the intermediate slices were automatically recognized and segmented by the software. To appropriately visualize the three-dimensional data, NVIDIA IndeX (NVIDIA, Santa Clara, US) and Avizo (Thermo Fisher Scientific, Waltham, US) was used, as previously described in Reichmann et al. (2023).

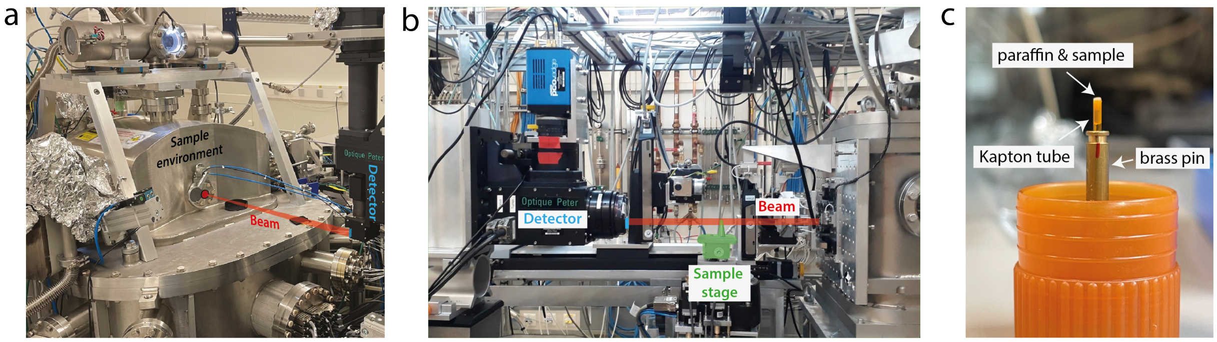


Figure 2: **a** Photograph depicting the SR2 setup at the ID16A beamline (ESRF, Grenoble, France). The sample is placed by a manually controlled gripper inside the sample environment in vacuum. After the exiting window the beam propagates through air before it hits the detector. **b** Photograph depicting the SR1 parallel beam setup at the P10 beamline (PETRA III, Hamburg, Germany). The monochromatic beam is attenuated by polished single-crystal attenuators (Si) and thereafter directly passes through the sample without any intermediate optical elements. **c** Sample as prepared for the experiment, biopsy surrounded by a Kapton foil inside Huber (brass) pin.

.

*Electron Density Calculations*

After phase retrieval and reconstruction, the electron density was calculated from the phase shift per unit length $\omega$ (reconstruction output), according to

$$\rho_e = \rho_{e,ref} - (\Delta\omega \frac{\lambda}{2\pi}) \cdot \frac{2\pi}{\lambda^2\ r_0} = \rho_{e,ref} - \frac{\Delta\omega}{r_0\lambda}, \tag{5}$$

where $\Delta\omega$ refers to the difference with respect to a reference value

$$\Delta\omega = \omega - \omega_{ref} = -\frac{2\pi}{\lambda}(\delta - \delta_{ref}), \tag{6}$$

and $\omega$ ($\omega_{ref}$) and $\delta$ ($\delta_{ref}$) denote the phase shift per unit length and the index of refraction decrement for the tissue of interest (paraffin), respectively (Henke et al. (1993)). $\rho_e$ is the electron density and $r_0$ is the classical electron radius ($\approx 2.818 \cdot 10^{-6}$ nm). A two-dimensional slice with the phase shift per unit length for each pixel is shown in Fig.3**a** (left), as well as the calculated electron density (Fig. 3**a**, right). The same ROI with the original gray values is shown in Fig. 6 (**d-f**).

For an estimate of the spatial resolution, Fourier-shell-correlation (FSC) van Heel & Schatz (2005) was performed. To this end, a scan with 4000 acquired projections was divided into two equally sized datasets of 2000 projections each (even/odd) and separately reconstructed. FSC was then evaluated as a function of the spatial frequency (in units of $\frac{2\pi}{px}$ on a region-of-interest ($1000^3$ pixel), selected from the center of the volume as implemented in Lohse et al. (2020). The half-bit threshold was used as the criterion for the spatial resolution estimation with the according threshold curve plotted in Figure 3**b** alongside the FSC-curve. The intercept, which determines the maximum resolvable spatial frequency, was found at 0.8 $\frac{2\pi}{px}$, yielding a half-bit resolution of 355 nm. Since the dataset was split, this value is a conservative estimate.

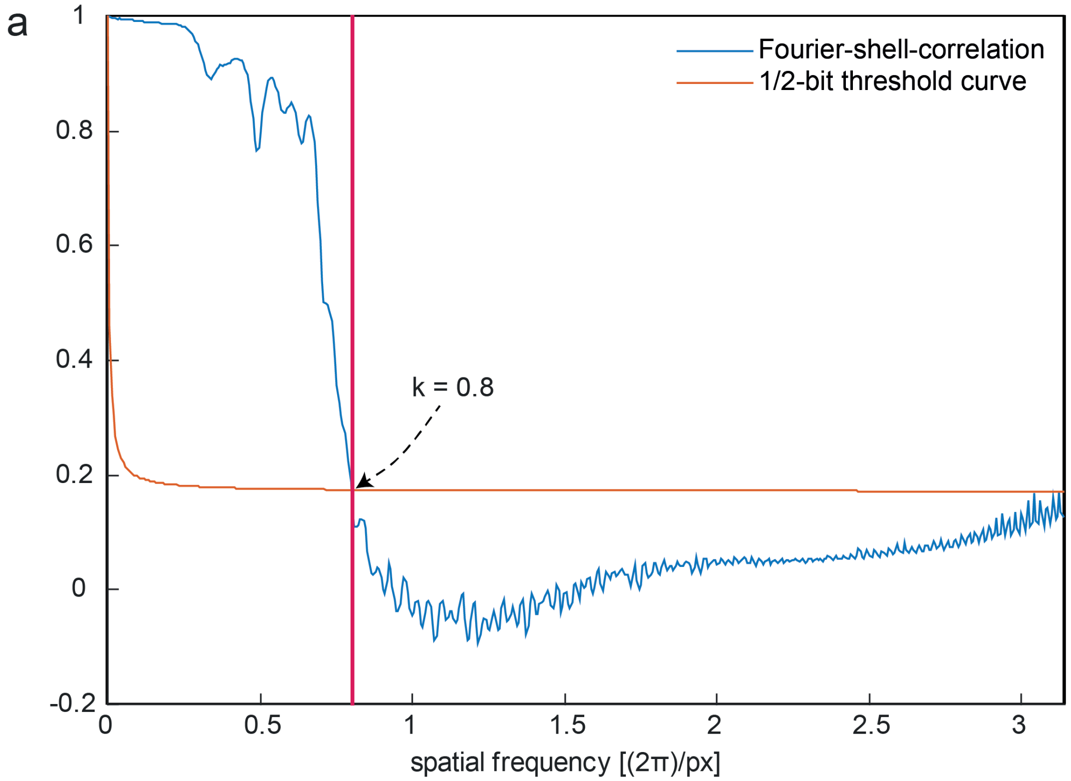


Figure 3: **a** FSC-curve plotted alongside half-bit threshold curve for a high resolution dataset acquired with the SR2-setup (Hirano-body dataset). Intercept at $k$ =0.8 $\frac{2\pi}{px}$ yields a half-bit resolution of 355 nm.

### Additional Data Analysis

#### *Correlation of XPCT recorded tomographic reconstruction to histological sections*

The procedure of a correlative comparison between different modalities, in this case stained histological slices investigated under a light microscope and slices from reconstructed XPCT-recorded volumes, is exemplified on a sample with an expected considerable amount of $A\beta$-plaques. After data acquisition by XPCT, the remaining 1 mm diameter paraffin biopsy is embedded back into an entire paraffin block. With guidance by the reconstructed volume, as common practice in histopathology, the tissue is cut with a microtrome in 3 $\mu m$ slices. These slices are then stained (H&E (Figure 4**a**, 6E10 (Figure 4**b**) or other antibody stains) and the slice is digitalized. In a next step, the

XPCT volume is browsed until certain features are spotted, visible in the histological slice. In this example, an elongated region was discovered where the sample matched almost perfectly (Figure 4**d,e**). Finally, the plane in the XPCT is rotated around the elongated axis previously found, until the other features match. Since the histological slice is not isotropic and has a thickness of several $\mu$m, finding a slice where all features perfectly match is strictly speaking impossible. While in Figure 4**c** any morphological features in the bottom and left part align well, this is not the case for the upper right part of the slice.

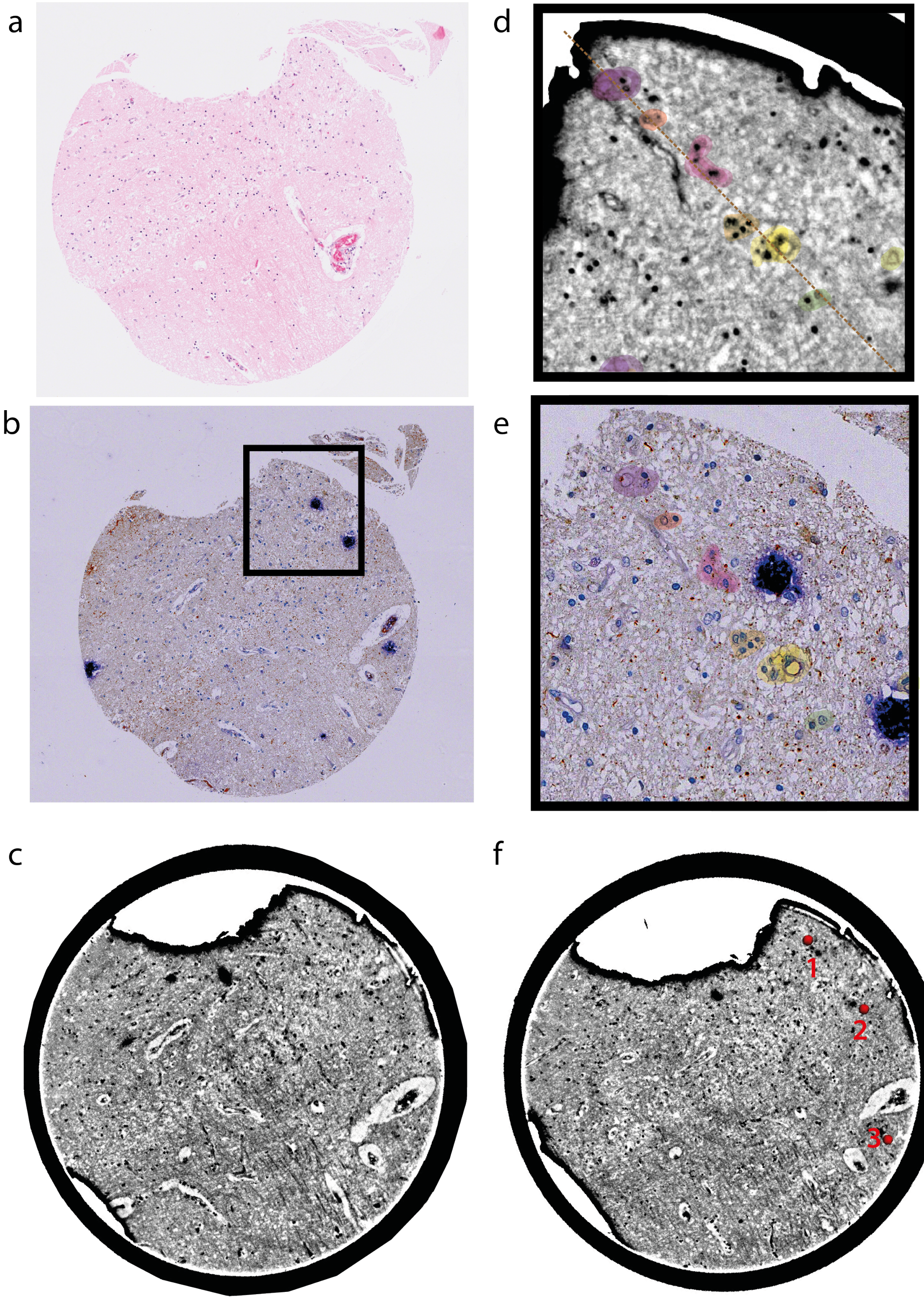


Figure 4: H&E stained (**a**) and 6E10 stained (**b**) slice from human hippocampal tissue (CA1) with a zoom into a plaque-rich area (**e**) and correlative slice in the XPCT dataset (**d**) recorded at the LS1 setup. (**c**) shows slice through 3D volume, where most features are aligned with the histological slice, while with further tilting the plaques become visible (**f**).

95 By further tilting the slice, the plaques finally come into the FOV (Figure 4**f**).

While a high-resolution slice from the SR2-setup would make a good complement, unfortunately the nano-imaging dataset starts approximately 400 slices (36 $\mu$m) above the histologically investigated slice. With an equivalent pipeline, data acquired of samples with Hirano bodies and vascular accumulation of $A\beta$ is compared to histological slices, confirming their presence with the respective light-microscopy stains.

101 *GVD and LB samples recorded at the SR1-beamline*

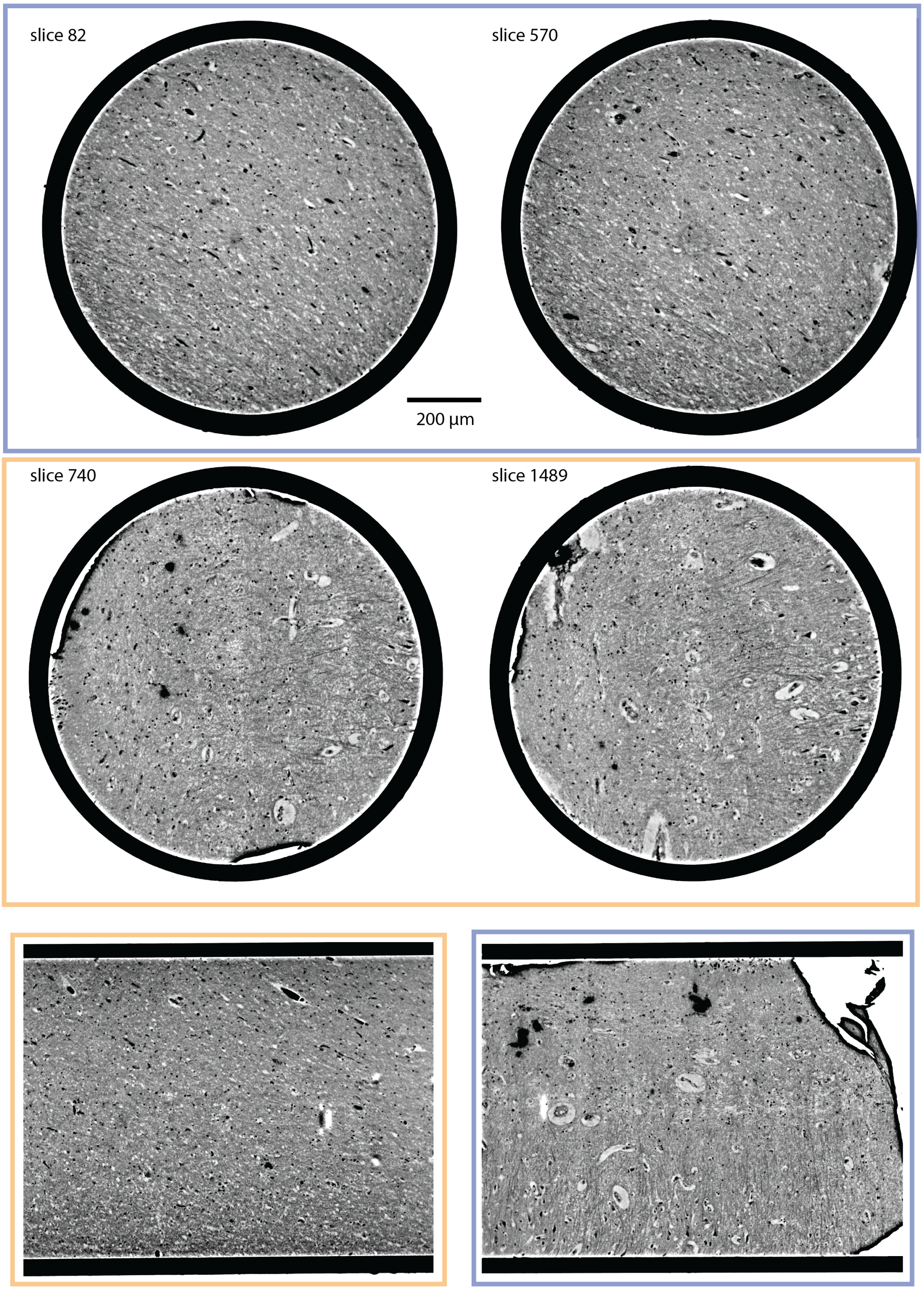


Figure 5: Additional xy-slices at different planes and orthogonal slices to GVD (orange) and LB (blue) samples recorded at the SR1-beamline.

*Correlation between high-resolution dataset recorded at SR2-setup and overview scan recorded at SR1-setup*

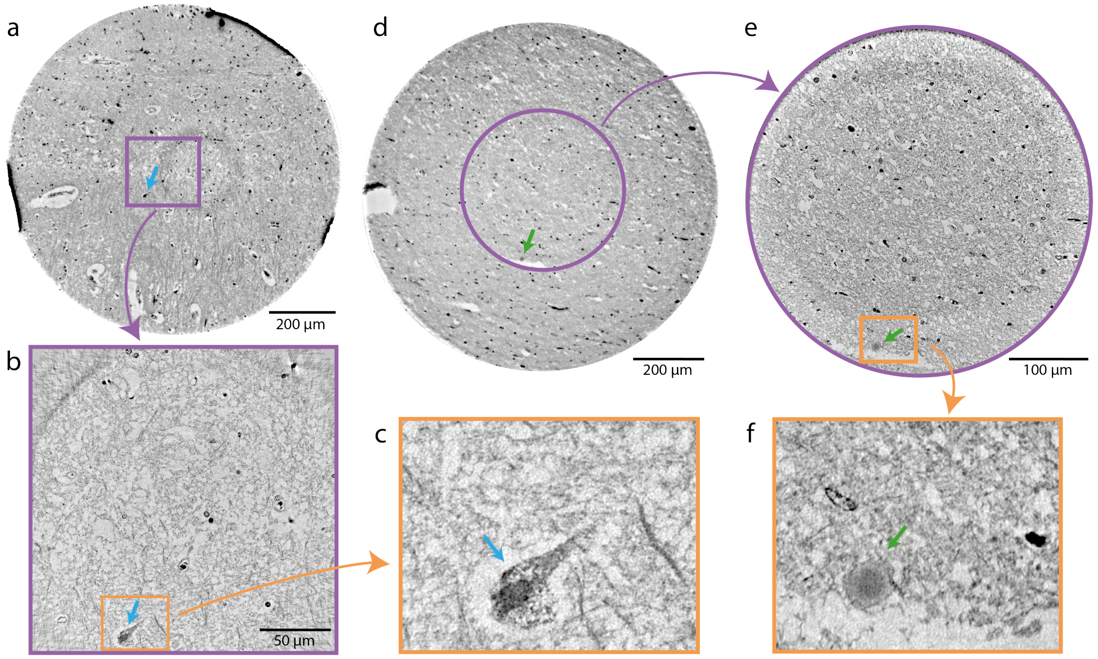


Figure 6: Comparison of data acquired at the parallel beam SR1 setup and the high-resolution SR2 setup. **a** Slice through reconstructed electron density of a 1 mm brain punch from patient with granulovacuolar degeneration (blue arrow) acquired at SR1 setup. **b** Same region as in **a**, recorded with high resolution SR2 setup with (**c**) zoom at granulovacuolar degeneration. **d** Slice through reconstructed electron density of a 1 mm brain punch from patient with Lewy bodies (green arrow) acquired at SR1 setup. **e** Same region as in **d**, recorded with high resolution SR2 setup with (**f**) zoom at a Lewy Body.

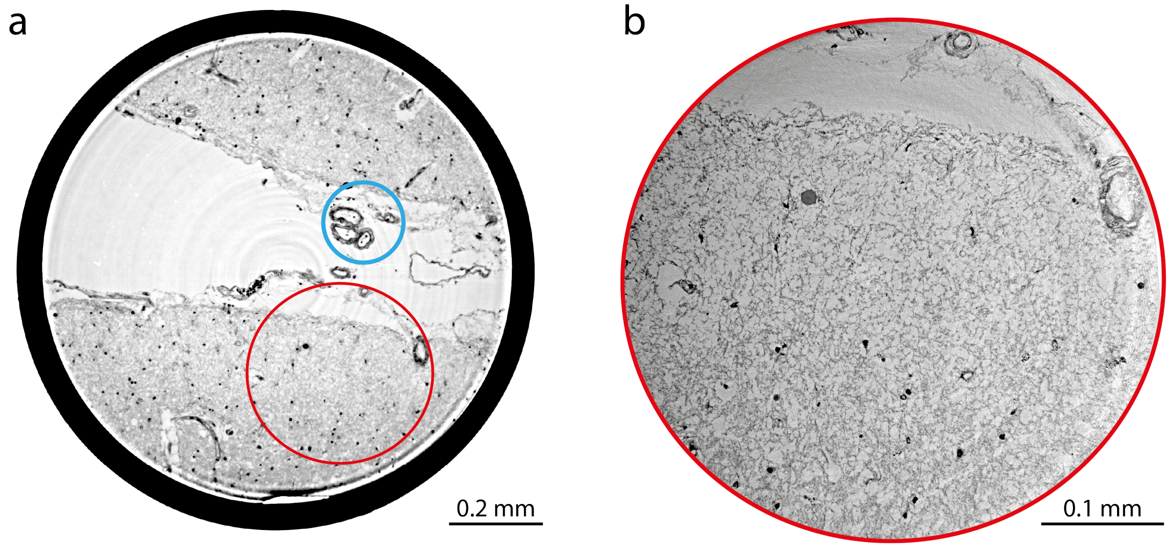


Figure 7: Vascular amyloid deposition in cerebral amyloid angiopathy (CAA). **a** Virtual section covering the entire 1 mm diameter of the punch biopsy (SR1-setup). **b** Section through a subset of the volume covered by the high resolution scan (SR2-setup).

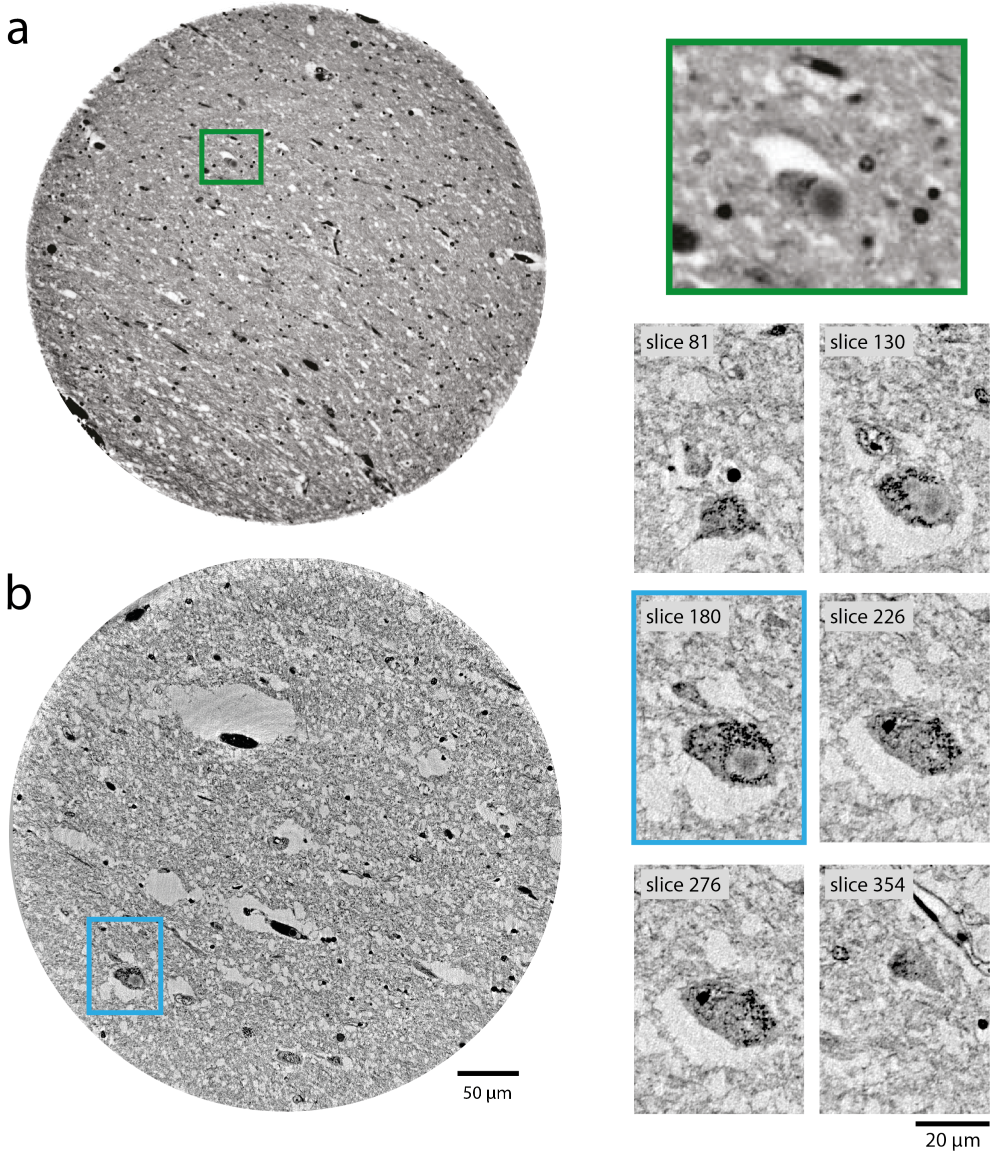


Figure 8: M. Parkinson. **a** Virtual section through the reconstructed volume of substantia nigra, recorded with the parallel beam setup (SR1, left), with a zoomed depiction of a Lewy body (right). **b** Reconstruction of the same sample from a high resolution scan recorded with the cone beam setup (SR2, left), and corresponding virtual sections through a Lewy body (right).

*Correlation between stained histological slices and according slice in overview scan recorded at SR1-setup*

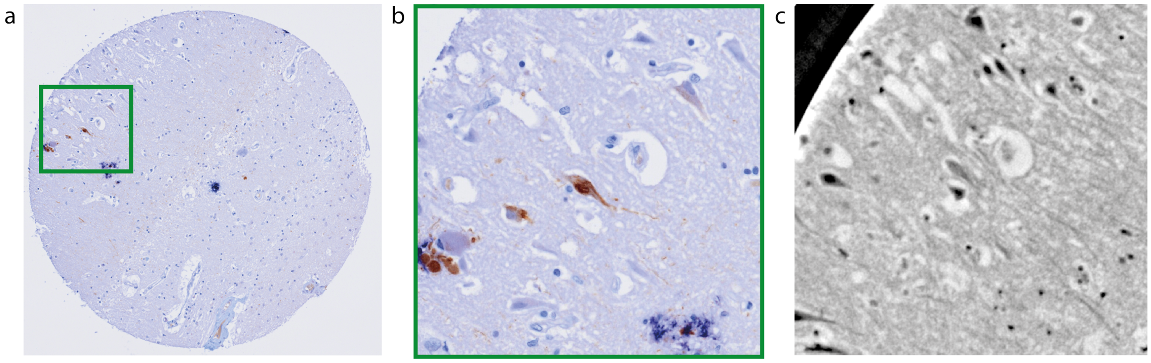


Figure 9: **a** Histological section in the hippocampal CA1 region of a brain sample with a NFT-specific stain (AT8) for intraneuronal neurofibrillary tangles (brown, visible in **b**). **c** Approximately equally oriented slice from XPCT scan. At the given resolution NFTs cannot be detected in the X-ray reconstruction. This may be possible at higher resolution, such as SR2.

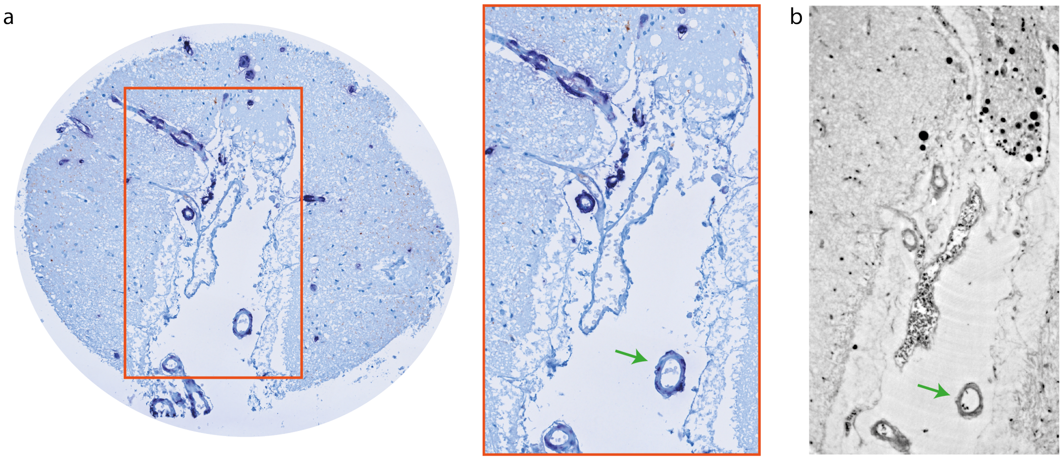


Figure 10: **a** Histological section in the hippocampal CA1 region of a brain sample with an $A\beta$-specific stain (dark blue). **b** Approximately equally oriented slice from XPCT scan.

*Additional visualization of Hirano body and granulovacuolar degeneration tomographic data*

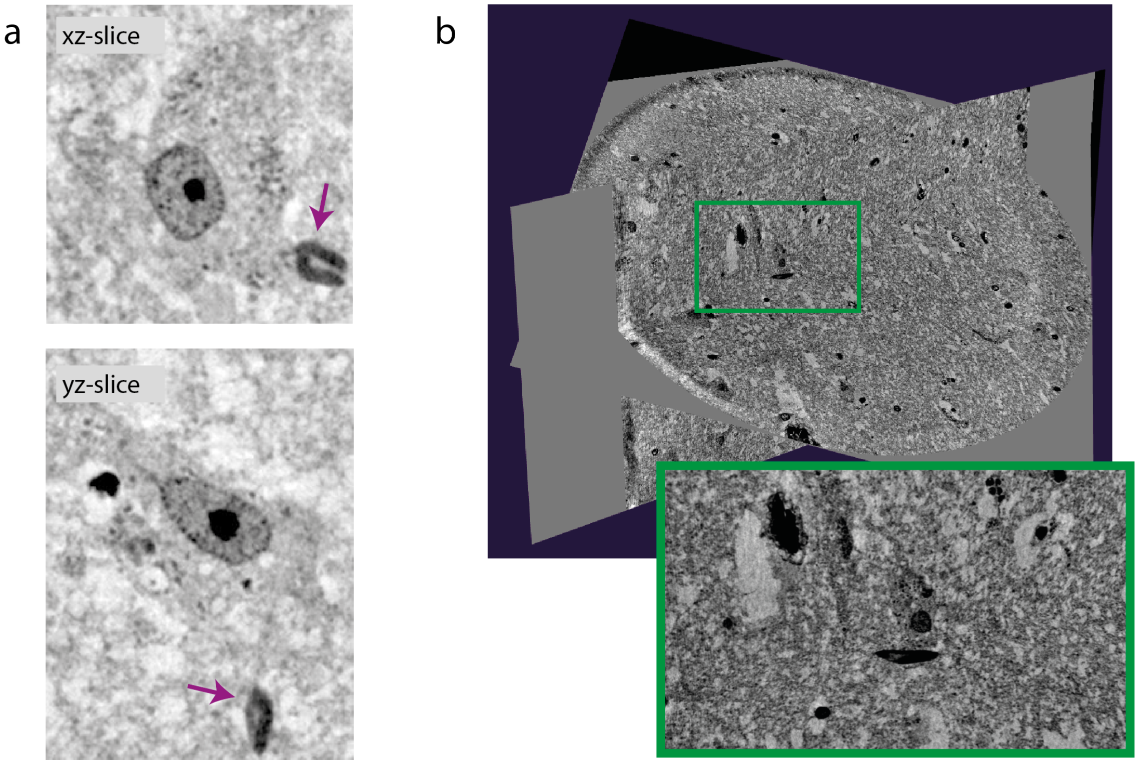


Figure 11: **a** Orthogonal sections through the same Hirano body as shown in main manuscript. **b** Differently oriented slices through three dimensional volume (visualization with viewer by Histomography GmbH).

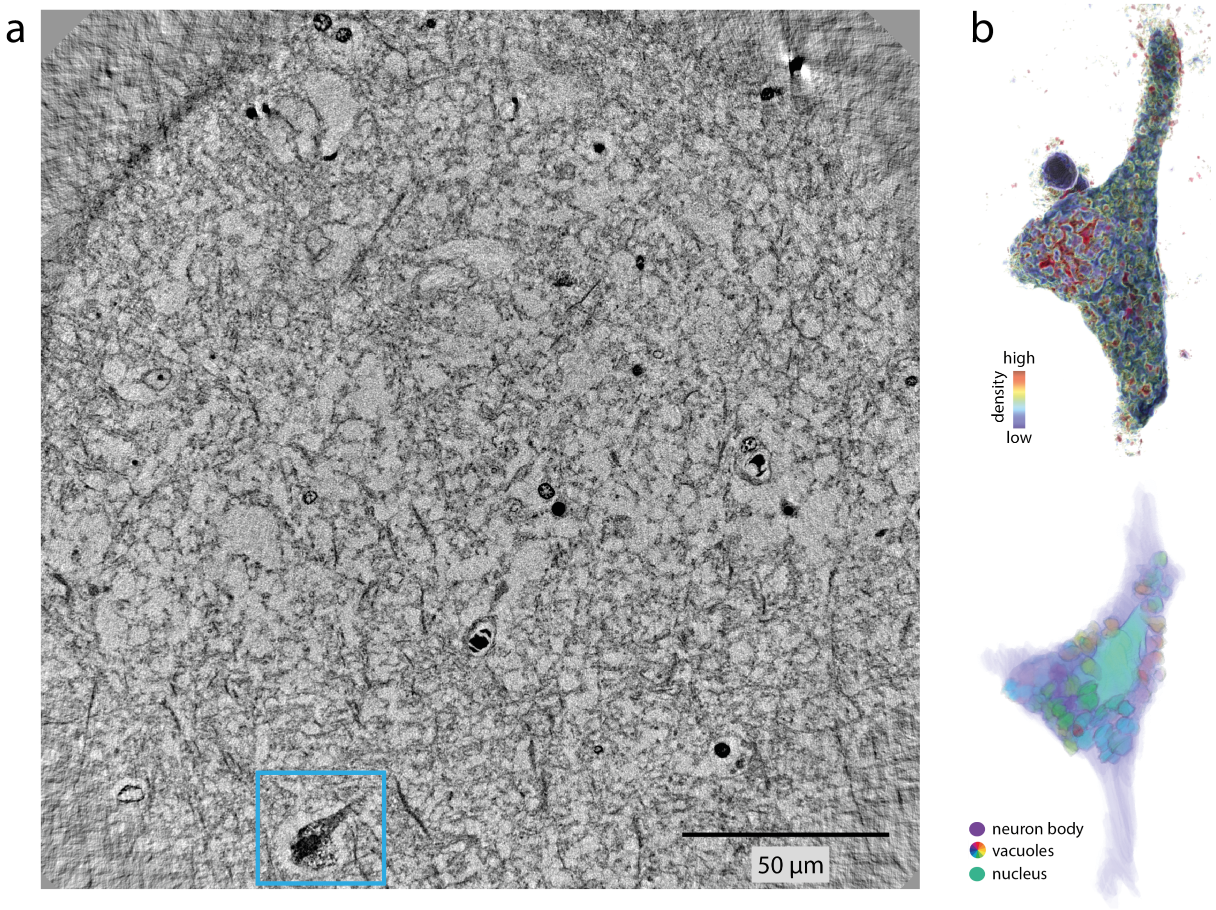


Figure 12: Granulovacuolar degeneration. **a** Virtual section through tomographic reconstruction of a scan from human hippocampus (CA1 region) with a neuron exhibiting GVD, characterized by vacuoles with a dark granule in the center (bottom left). **b** Rendering of the degenerating neuron (top, NVIDIA IndeX) and segmentation of separate vacuoles (each in different color) with cell nucleus (green) and the neuron body (violet).
